# Modeling spillover dynamics: understanding emerging pathogens of public health concern

**DOI:** 10.1101/2023.10.02.23296428

**Authors:** Fernando Saldaña, Nico Stollenwerk, Joseba Bidaurrazaga Van-Dierdonck, Maíra Aguiar

**Affiliations:** Basque Center for Applied Mathematics (BCAM), Bilbao, Spain; Ikerbasque, Basque Foundation for Science, Bilbao, Spain; Public Health, Basque Health Department, Bilbao, Spain; Dipartimento di Matematica, Università degli Studi di Trento, Italy

## Abstract

The emergence of infectious diseases with pandemic potential is a major public health threat worldwide. According to the World Health Organization, around 60% of the reported emerging infectious diseases are zoonoses and have been triggered by spillover events. Although the dynamics of spillover events are not yet well understood, mathematical modeling has the potential to characterize the highly complex interactions among pathogens, wildlife, humans, and the environment where they coexist. In this work, motivated by discussions on the introductory phase of SARS-CoV-2 towards a pandemic scenario, we address the so far unexplored the emergence of novel infectious agents. Aiming at gaining insights into the dynamics of spillover events and the final outcome of an eventual disease outbreak in a population, we propose a continuous time stochastic modeling framework to describe a cross-species disease transmission by coupling the dynamics of animal reservoirs and human hosts. A complete analysis of the system is conducted, followed by numerical experiments where we investigate different scenarios of spillover events. Applied to the emergence of SARS-CoV-2 and monkeypox viruses, our results provide insights into the emergence of new infectious diseases able to cause not only local outbreaks but eventually explosive epidemics towards a pandemic.

## 1 Introduction

The COVID-19 health crisis has been a stark reminder of the vulnerability of the human population to novel pathogens and a call for global action to reduce the risk of future emerging infectious diseases (EIDs)^1^. But SARS-CoV-2 was not the first and certainly not the last pathogen to jump across species from wildlife to the human population in a zoonotic spillover event. Even before the emergence of SARS-CoV-2, it was estimated that zoonotic pathogens brought approximately one billion human infections and millions of deaths annually^2^. Moreover, due to increasing levels of contact between humans and wildlife, the frequency of spillover events seems to be higher than ever before^3^. The unprecedented monkeypox outbreaks that arose outside endemic regions during the summer of 2022 is another example of how public health threats are enhanced by the movement of individuals worldwide^4^. With clear evidence of how devastating an emergent zoonotic pathogen can be, it is crucial to strengthen the public health capacity for detecting, identifying, and addressing likely explosive epidemics.

Spillover events depend on highly complex interactions among several factors, including the prevalence and intensity of the infection in the natural reservoir, pathogen features that determine its ability to be transmitted between different hosts, and population characteristics that affect the probability of infection^5^. Recent evidence suggests that changes in land use, such as deforestation and intensification of agriculture, have increased the probability of the emergence of zoonotic diseases^6^. The sale and consumption of wild animal species can also increase the risk of zoonotic spillover^7^. Furthermore, high population density tied to high human mobility leads to a greater possibility that such spillover events lead to deadly pandemics^8^. Zoonotic epidemics have had devastating health and socioeconomic impacts with an estimated annual economic loss of US212 billion since 1918^3,6^. Yet, preventive actions towards the reduction of EIDs would be reasonably low-priced in comparison with the economic and mortality costs of responding to large epidemic outbreaks^6,9^.

Nowadays, it is well-recognized that human and animal health are deeply interconnected and linked with the environment where they coexist. Strategic mathematical modeling is vital for understanding this connection and therefore the ecology of future EIDs. Mathematical modeling also has the potential to improve disease surveillance, undertake dynamic risk assessments, and establish interventions for prevention and control. Nevertheless, zoonotic spillover events are rarely included in theoretical disease transmission models for EIDs^10^, and the dynamics of pathogen emergence causing a new infectious disease with pandemic potential remains largely unexplored. Major difficulty relies on the lack of quality data, and a number of research groups struggle with the current challenges of modeling epidemics. Most of the existing models focus on understanding the spread of already-established infectious agents within a single population, either the reservoir component or the human host^7,10^. Although a number of modeling studies have incorporated pathogen spillover transmission into multi-host epidemic models to investigate EIDs^11–20^ the stochastic nature of such events is often not fully investigated. While some recent studies have analyzed different aspects of the zoonotic spillover interface and have brought new and important insight on the emergence of new pathogens through spillover dynamics^14,16,18,21^, much remains to be understood and there are interesting avenues for future research^22^, including more comprehensive modeling frameworks to represent the complete evolution of the zoonotic spillover process; especially to capture disease dynamics in non-human species^10,17^. We propose a continuous time stochastic modeling framework to describe a cross-species disease system using a minimalist two-species epidemiological model. We consider three routes of transmission (i) animal-to-animal, (ii) animal-to-human, and (iii) human-to-human, so the two-host populations are fully coupled. While we neglect the possibility of spillback events i.e. human-to-animal transmission, we evaluate different progressive stages of zoonotic emergence. Our modeling framework uses data on laboratory-confirmed cases to investigate the 2022 monkeypox outbreaks outside the endemic region, with further discussions on the initial phase of the COVID-19 pandemic. Studying the coupled epidemic in our two-species model allows us to investigate the role of the reservoir and the spillover transmission rate on the emergence and size of the outbreaks in the human population, gaining insights into the impact of EIDs risks in the public health context.

## 2 Results

Spillover events are significantly affected by the epidemiological dynamics in the reservoir, such as prevalence, distribution, and intensity of infection. Initially, pathogens exist within their ‘natural’ reservoir, which can be simply defined as an animal population that can maintain a pathogen in circulation^23^. As a consequence of this definition, the basic reproduction number in the reservoir, 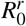, must be greater than one. Due to a long history of host–pathogen co-evolution, the natural animal host usually carries pathogens without presenting any clinical symptoms (or at most mild clinical symptoms not affecting their health). For example, some of the most important zoonotic viruses such as HIV, Ebola virus, coronaviruses, influenza A viruses, hantaviruses, or henipaviruses, can result in severe disease in humans; yet, the reservoir hosts usually tolerate these infections with little or no overt of disease^24^.

With that in mind, for the reservoir dynamics, we propose a simple SI (susceptible-infected) model coupled with a SHAR (susceptible-hospitalized-asymptomatic-recovered) model for the human host, as described in Section 4. By assuming that animal hosts are natural carriers of the pathogen, disease transmission dynamics is in stationarity, with the basic reproduction number in the reservoir set to the supercritical regime, i.e. above one (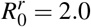, with the average reservoir life expectancy, assumed to be 1*/d*_*r*_ = 1 year, and the transmission rate *β*_*r*_ = 2*d*_*r*_). The numerical simulations in Figure 1 show the stochastic realizations and the analytical mean-field solution for the infected reservoir population in the coupled SI-SHAR model.

**Figure 1.**
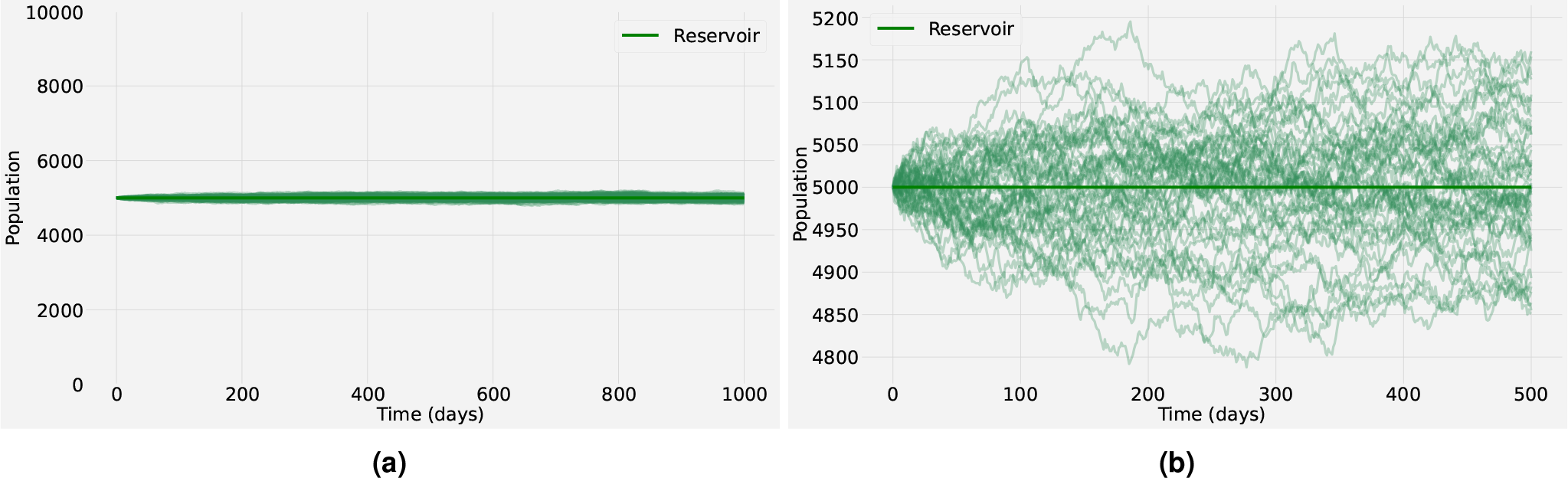
Stochastic realizations and the analytical mean-field solution for the infected reservoir population. The baseline values of the model parameters are shown in Table 1 in the SM. (a) shows the full spectrum for the infected reservoir population, whereas (b) provides a closer look into the stochastic fluctuations (thin lines) observed around the endemic equilibrium (bold line). The baseline parameter values used in these computations are listed in Table 1 in the SM.

### 2.1 The trade-off between the human basic reproduction number and the spillover rate

In a deterministic setting, a spillover event will successfully establish an epidemic in the human population if the basic reproduction number satisfies *R*_0_ *>* 1 (super-critical regime), whereas no self-sustained transmission can occur otherwise. These mean-field dynamics are usually valid if a sufficiently large number of infected individuals are present to make statistical variations negligible. However, stochastic systems display large fluctuations resembling new epidemic waves even in the sub-critical regime (*R*_0_ *<* 1). Nevertheless, in the super-critical regime (*R*_0_ *>* 1), many realizations maintain low numbers of infections, needing a long time to take off and become an epidemic wave. These fluctuations, rather than details of the infection process, dominate the dynamic behavior of the system. In particular, under the homogeneous mixing assumption, for an initial seed of *n* infected individuals, the probability of extinction is 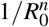. Correspondingly, the probability of a major outbreak is 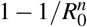. These estimates are valid for several Kermack-McKendrick-type models^25^

Building on the work of Wolfe et al.^26^, Lloyd-Smith et al. used the human reproduction number 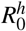 to propose a classification scheme of pathogen emergence^27^. In this framework, pathogen emergence is divided into five progressive stages ranging from agents that exclusively infect animals (Stage I) to those that only infect humans (Stage V)^26,27^. Zoonotic emergence takes place in stages II-IV. In stage II, spillover events only lead to primary infections in humans but no further human-to-human secondary transmission 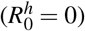 occurs. In stage III, pathogens can lead to limited transmission chains of human-to-human secondary infections that stutter to extinction 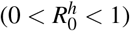, while in Stage IV pathogens still arise in animal populations and can be successfully transmitted between human hosts, leading to self-sustained chains of transmission 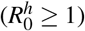^26,27^. Although this classification scheme is helpful, it is based solely on the basic reproduction number for the human population, and the role of the natural reservoir and spillover events on the epidemiological dynamics is not discussed^19^. In a deterministic setting, we have shown (see Supplementary Material) that for the coupled SI-SHAR model (5), if the reservoir is at the endemic equilibrium and the spillover rate satisfies *τ >* 0, then no disease-free equilibrium (DFE) exists for the human population. In other words, if there is a recurrent transmission from the natural reservoir to humans, the deterministic process always predicts an outbreak in the human population. Hence, the deterministic model lacks the capability to properly simulate important dynamics of pathogen emergence at crucial regimes such as the introductory phase where limited transmission chains of human-to-human mostly end up with disease extinction. Within the stochastic framework, the occurrence of a disease outbreak in the human population will depend mainly on 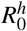 and the spillover rate *τ*. We provide numerical experiments to illustrate this trade-off. For the numerical simulations we assume that at the initial time, the human population is fully susceptible. The infection prevalence level is considered close to the endemic equilibrium in the reservoir (see Figure 1). Note that non-endemic prevalence levels in the reservoir do not change the qualitative behavior of the long-term human epidemiological dynamics as long as the reservoir converges to its endemic equilibrium (see Supplementary Material). The disease severity ratio is fixed as *η* = 0.2 so 20% of the population develops severe symptoms prone to hospitalization upon infection. First, it is assumed that natural immunity after recovering from infection is lifelong; yet, the possibility of a loss of immunity is also considered in this study.

Quantifying the rates at which novel zoonotic pathogens spill over into human populations is a challenging task for public health officers. Even for already circulating pathogens such as COVID-19, monkeypox or Ebola viruses, detection can be problematic in areas with limited access to confirmatory diagnostics. Hence, a reliable estimation of the spillover force of infection is extremely difficult to obtain^27^. Yet, subtle changes in the spillover rate might lead to substantial changes in the human epidemiological dynamics^14^. Figure 2 shows a typical stochastic realization for the total human disease prevalence (adding the asymptomatic and hospitalized classes) for different values of the pathogen spillover rate (*τ* ∈ [10^−6^, 10^−3^]). The dynamics correspond to stage II of pathogen emergence, with *β* = 0 and 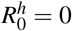 so the outbreak in the human population is fully driven by spillover events and no further human-to-human secondary transmission is possible. Observe that for *τ* = 10^−6^ (see Figure 2a) the prevalence never surpasses one active human infection and there can be a huge time gap between consecutive spillover events. For *τ* = 10^−5^ or *τ* = 10^−4^ (see Figures 2b and 2c, respectively), while the prevalence levels rise, the increment on the number of cases is not significant because the maximum prevalence is even below 0.1% of the total population. Furthermore, even if the time gap between consecutive spillover events decreases there are still periods with zero human infections (see Figures 2b and 2c). By contrast, for *τ* = 10^−3^ human prevalence levels are always positive, resembling the supercritical regime of the dynamics (see Figure 2d).

**Figure 2.**
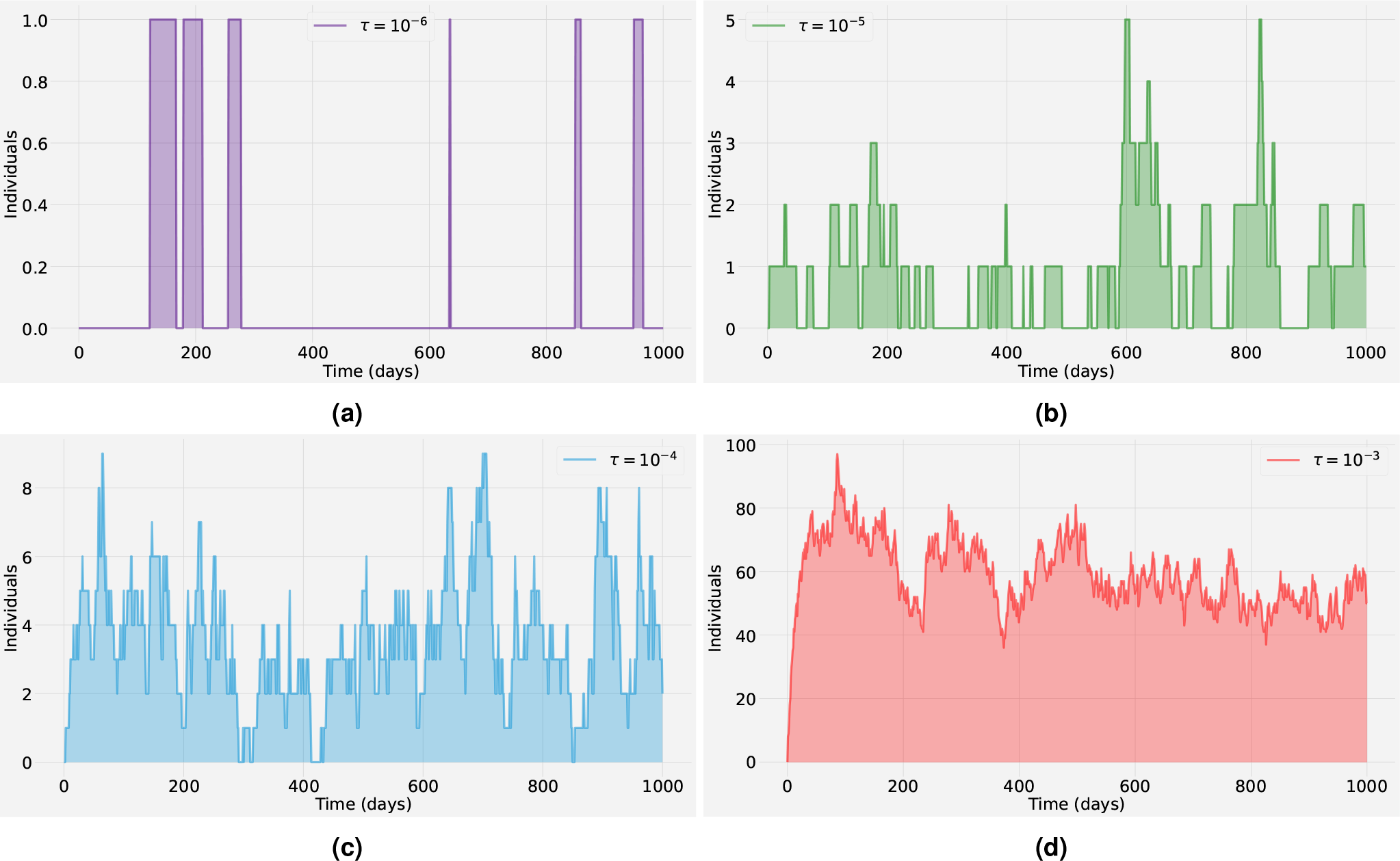
A typical stochastic realization for the dynamics of the infection prevalence (adding the asymptomatic *A* and hospitalized *H* classes) in a human population. For different spillover rates (a) *τ* = 10^*-*3^, (b) *τ* = 10^*-*4^, (c) *τ* = 10^*-*5^, (d) *τ* = 10^*-*6^, the outbreak in the human population is fully driven by spillover events (with *β* = 0 and 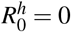). The baseline parameter values used in these computations are listed in Table 1 in the SM.

Figure 3 presents an ensemble of stochastic realizations that correspond to stage III of pathogen emergence in a human population. In this stage, human-to-human secondary infections are possible but without further spillover events, they cannot lead by themselves to a large epidemic outbreak in the human population^27^. Therefore the basic reproduction number in the human population 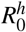 is less than one. In other words, newly infected individuals might generate new infections before recovery but on average they recover faster than they generate new infections, thus after a small outbreak the system is expected to end with almost zero cases on average (eventually with large excursions towards high outbreaks)^28^. In Figure 3 the blue thin lines correspond to stochastic realizations for the asymptomatic class, whereas the orange thin lines correspond to the hospitalized class. The respective bold lines represent the mean-field solutions. It is assumed that 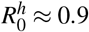. Observe that for a spillover rate *τ* = 10^−5^ (see Figure 3a) many very small outbreaks occur but none of them lead to an epidemic breakout. Whereas, for a higher spillover rate *τ* = 10^−4^ (see Figure 3b), prevalence levels appear to undergo a short but clear exponential growth of infections that seems like a classical epidemic wave, even though the system is in the subcritical regime.

**Figure 3.**
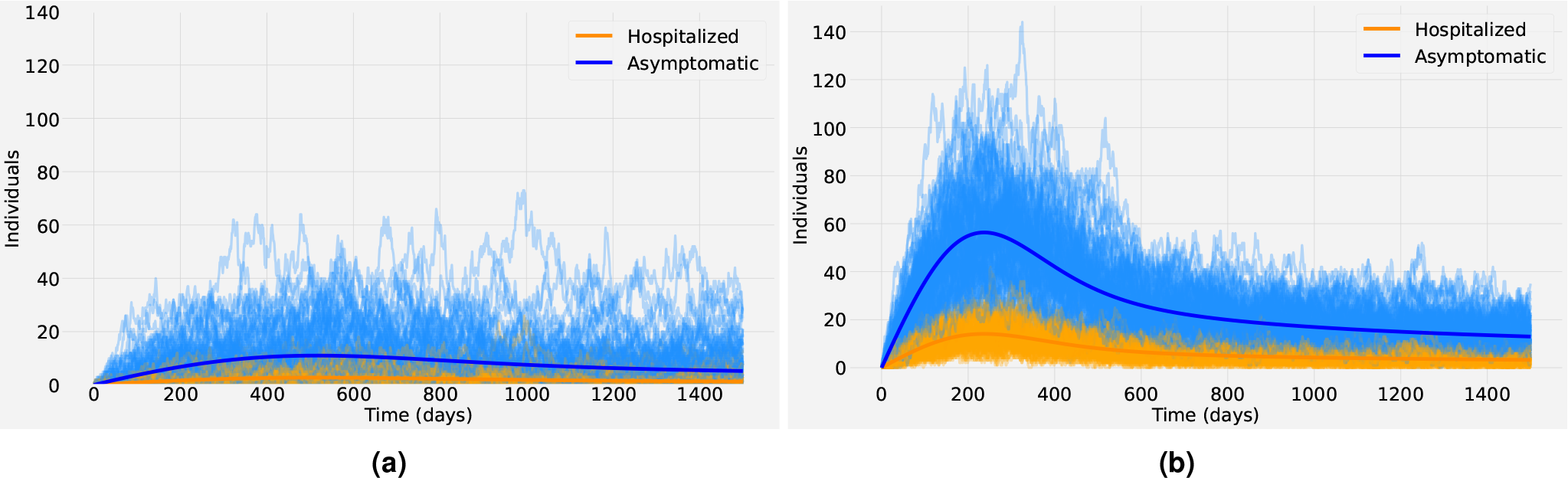
Stochastic realizations (bold lines) and the analytic mean-field solution (thin lines) for the asymptomatic *A* class (blue) and the hospitalized *H* class (orange). In (a) the spillover rate is *τ* = 10^*-*5^ and in (b) is *τ* = 10^*-*4^. The dynamics correspond to stage III of pathogen emergence with 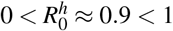, close to criticality. The baseline parameter values used in these computations are listed in Table 1 in the SM.

One key result that comes from comparing Figures 2d and 3a is that qualitatively the epidemiological dynamics in the infected human population are almost the same, even though in Figure 2d 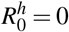 with *τ* = 10^−3^ while in Figure 3a 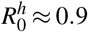 with *τ* = 10^−5^. Hence, if the human epidemic is within a sub-critical regime, spillover events can significantly affect the human host’s incidence patterns and prevalence levels. This finding is consistent with results in previous studies which found that besides stage IV, stages II-III can also lead to large outbreaks in the human population if driven sufficiently hard by the reservoir^18^. An important challenge when analyzing zoonotic outbreaks with short chains of transmission is thus to quantify the relative contribution of spillover events and human-to-human transmission to the force of infection. Mathematically, both spillover and self-sustained human transmission can be conceptualized as arrival processes^7,14^. Likely, the most significant difference is that spillover events are a series of one-time events that are commonly assumed to arise from random and independent contacts between the animal reservoir and the human host. Hence, the accumulation of spillover events is expected to grow linearly as a function of time. On the other hand, onward human-to-human transmission is better described by self-exciting events that can lead to short transmission chains or epidemic breakout with early exponential growth^7,14^.

The first row in Figure 4 shows the epidemiological dynamics in the human population that correspond to stage IV of pathogen emergence with 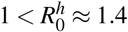 for *τ* = 10^−5^ (Figure 4a), *τ* = 10^−4^ (Figure 4b) and *τ* = 10^−3^ (Figure 4c). Observe that the size of the stochastic fluctuations of the infected cases increases as the value of the spillover rate decreases. For instance, in Figure 4a with *τ* = 10^−5^, we can observe that the peak time for some atypical stochastic realizations can be delayed up to 300 days in comparison with the peak time of the mean-field solution. These large deviations from the mean are not presented either in Figure 4b where *τ* = 10^−4^ or in Figure 4c where *τ* = 10^−3^. The second row in Figure 4 shows the peak time distribution based on an ensemble of 10^4^ stochastic realizations for the corresponding spillover rates. Robust estimation of the peak time distribution is crucial to understand the epidemic evolution and can help decision-makers to impose control policies. Applied to a real-world epidemiological scenario, note that even with significant advances in determining the magnitude and timing of epidemic curves, several epidemic peak time predictions for COVID-19 were inaccurate, with most peaks delayed with respect to predictions^28–31^. One explanation in this direction is that time-dependent changes in epidemiological parameters such as transmission and recovery rates have a non-trivial effect on the epidemic peak time^29^. Furthermore, it was found that imported cases, that is, individuals becoming infected by an undetected infection chain started outside the studied population, can significantly affect epidemic spread^28,32^.

**Figure 4.**
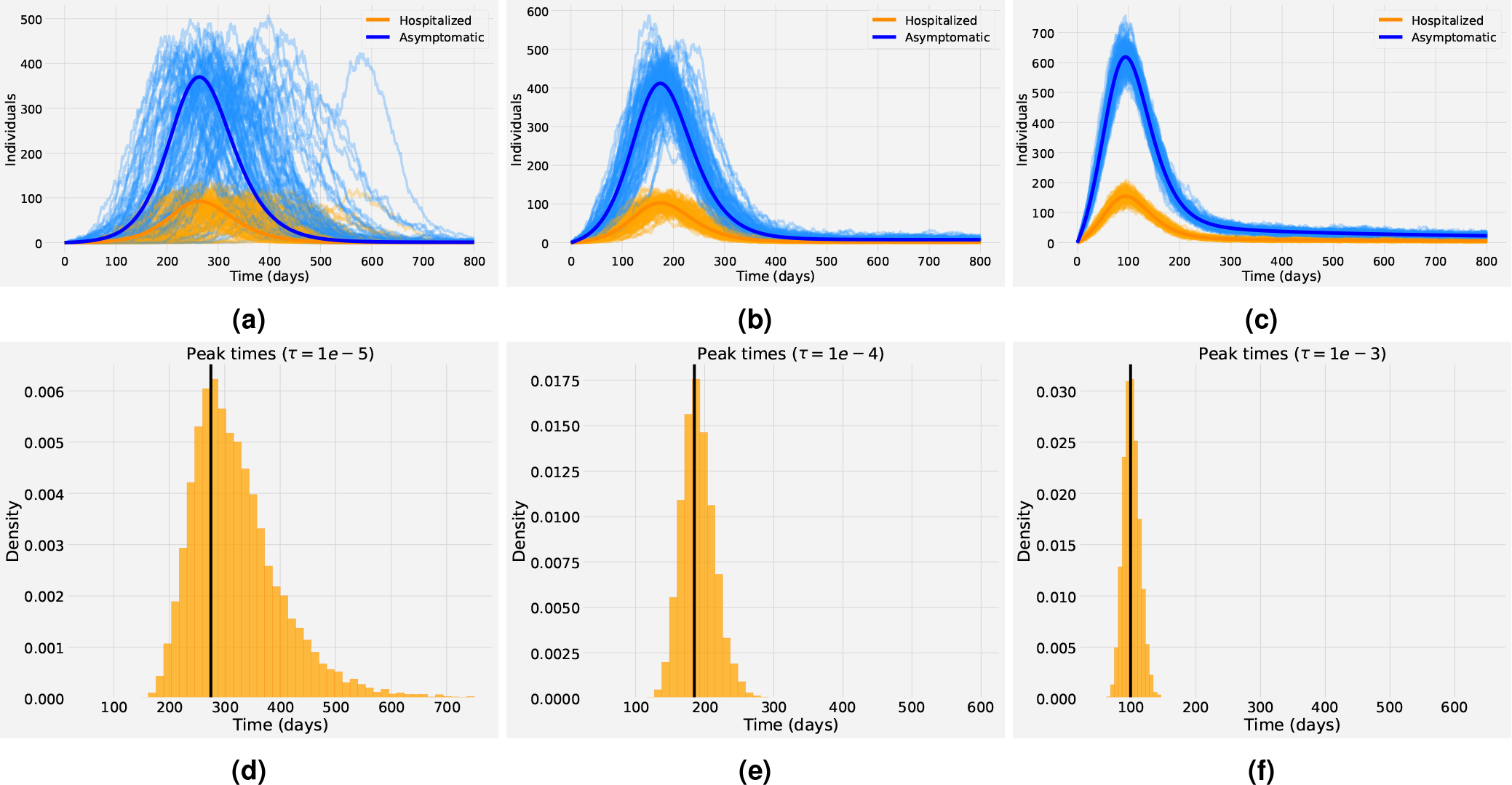
(First row): One hundred stochastic realizations and the analytic mean-field solution for the asymptomatic class (blue) and the hospitalized class (orange). Different spillover rates are investigated. In (a-d) *τ* = 10^*-*5^, in (b-e)*τ* = 10^*-*4^ and in (c-f) *τ* = 10^*-*3^. (Second Row): Peak time distribution for the corresponding stochastic realizations based on an ensemble of 10^4^ stochastic realizations. The x-axis corresponds to the time measured in days and the y-axis is the bin’s density. The vertical black line corresponds to the peak time of the outbreak obtained from the mean-field solution for each case, respectively. The dynamics correspond to stage IV of pathogen emergence with 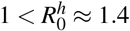. The baseline parameter values used in these computations are listed in Table 1 in the SM.

Applied to COVID-19 dynamics, Aguiar et al.^28,33,34^ have shown that models considering a small proportion of imported cases contributing to the overall force of infection can spark small or larger isolated outbreaks with growth factors and momentary reproduction ratios hovering around the epidemic threshold. The imported case was assumed to be a mobile asymptomatic infected individual, either a foreigner visiting the region or a local returning to the country without being detected by the current testing strategy, similar to what one expects when country lockdowns are completely lifted and mobility has returned to normal. Thus, the import factor refers to the possibility of susceptible individuals becoming infected by an undetected infection chain started outside the studied population, and can be directly related to the spillover transmission described here.

In Figure 4d one can observe that for a low spillover rate, most stochastic realizations reach the epidemic peak time after the mean-field solution, which explains why several COVID-19 peaks were delayed with respect to predictions. The distribution of the prevalence of the infection at different times (*t*_1_, *t*_2_, *t*_3_) for the three values of the spillover transmission rate is also investigated (see Figure 5). The time *t*_1_ is 50 days before the mean-field peak time, whereas *t*_2_ is at the day of the peak of the outbreak, and *t*_3_ is 50 days post-peak. The dynamics correspond to stage IV of pathogen emergence with 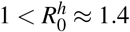. Observe that independent of the time chosen, for low levels in spillover transmission the prevalence distribution is highly non-Gaussian but as *τ* increases the distribution resembles more the bell-shaped form of the Gaussian distribution.

**Figure 5.**
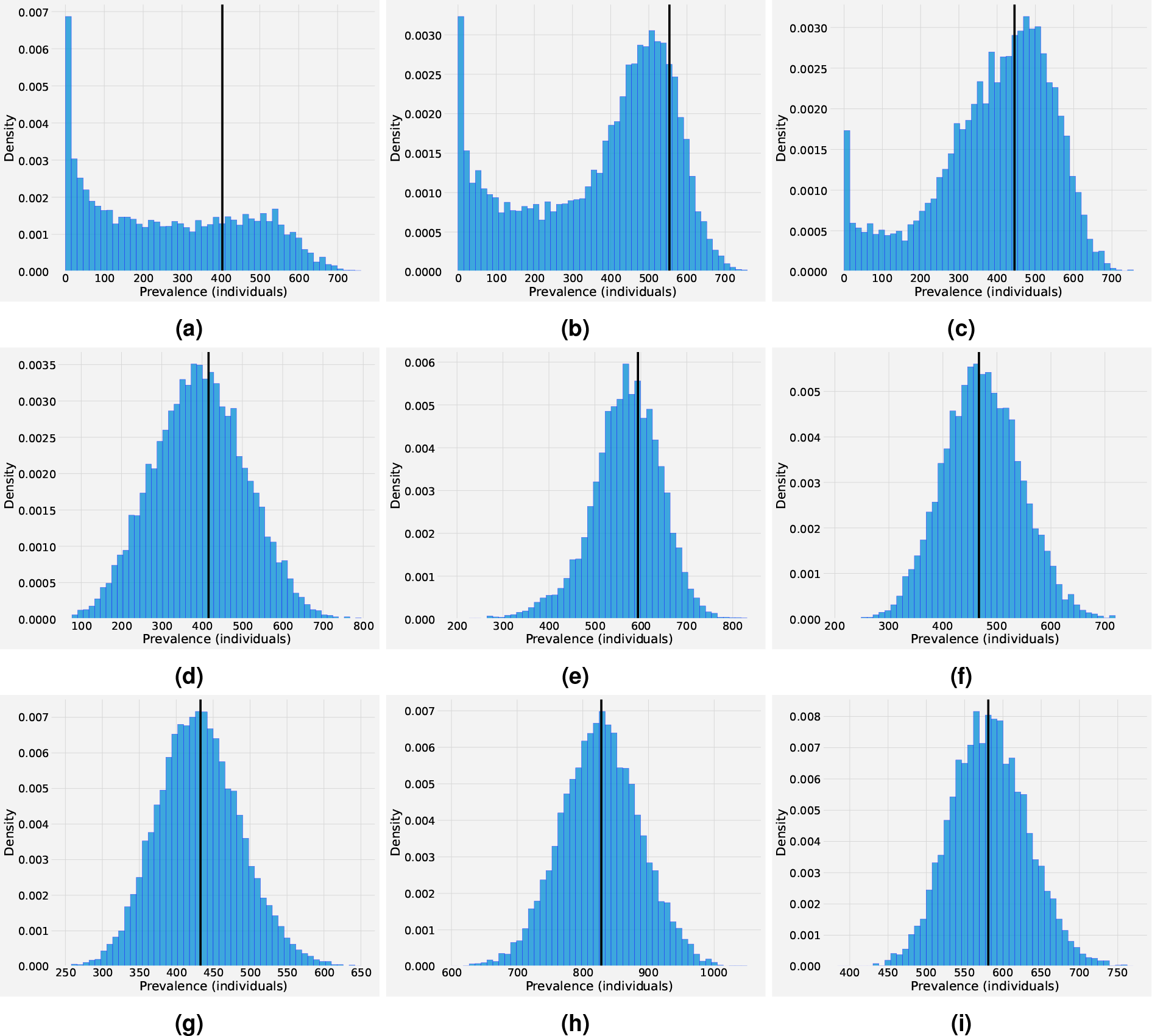
Distribution of the overall infected class, asymptomatic and hospitalizations (*H* + *A*), based on an ensemble of 10^4^ stochastic realizations for different peak time *t* considering different values of the spillover transmission rate. Left column: The time *t*_1_ is 50 days before the mean-field peak time. Middle column: The time *t*_2_ is the day of the peak. Right column: The time *t*_3_ is 50 days post-peak. Top row: The spillover transmission rate is *τ* = 10*e*^*-*5^. Middle row: The spillover transmission rate is *τ* = 10*e*^*-*4^. Bottom row: The spillover transmission rate is *τ* = 10*e*^*-*3^. The vertical black line corresponds to the mean field prevalence for each case, respectively. Other parameters used in these computations are listed in Table 1 in the SM.

In a deterministic setting, a clear separation exists between stage III and stage IV via the human basic reproduction number. However, when estimating epidemic risk at the early phase of epidemic outbreaks it can be challenging to distinguish between subcritical short chains of transmission and epidemic transmission with supercritical dynamics. One of the main challenges is to obtain a reliable estimation of the reproduction numbers. Although there are several non-parametric methods to key epidemiological parameters directly from early incidence data, recently emerged zoonoses usually come with data limitations that lead to highly uncertain estimates. Furthermore, due to stochastic effects, there is a chance of epidemic fade-out even if 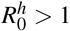 so only stage III dynamics are presented; whereas for the deterministic model, the infection establishes itself in the population and presents stage IV dynamics. Alexander et al. ^7^ notice this flaw in the classification scheme of Lloyd-Smith et al.^27^ and proposed an improvement considering an extra degree of freedom for stage IV that distinguish the probability of epidemic fade-outs and breakouts. A key issue to improve science-driven interventions for disease prevention and control is thus identifying the outbreak regime and the mechanisms that allow a stuttering chain to switch into a sustained transmission chain^28^.

### 2.2 Waning immunity and multiple epidemic waves

The interaction between pathogens and the host immune system is a highly dynamic process. In a few cases, the natural immune protection that develops after infection confers lifelong immunity. Nevertheless, the outcome of host-pathogen interactions is usually complex leading only to temporary or partial immune protection^35^. Figure 6 shows the effect of waning immunity and the possibility of reinfections. The average duration of natural immunity is assumed to be 180 days in Figure 6a and 360 days in Figure 6b. Figure 6 suggests that the loss of immunity in previously infected individuals replenishes the pool of susceptible individuals and might lead to seasonal epidemics or multiple epidemic waves. Such behavior has been observed in several local and nationwide COVID-19 epidemics since 2020^28,31,33,36–39^.

**Figure 6.**
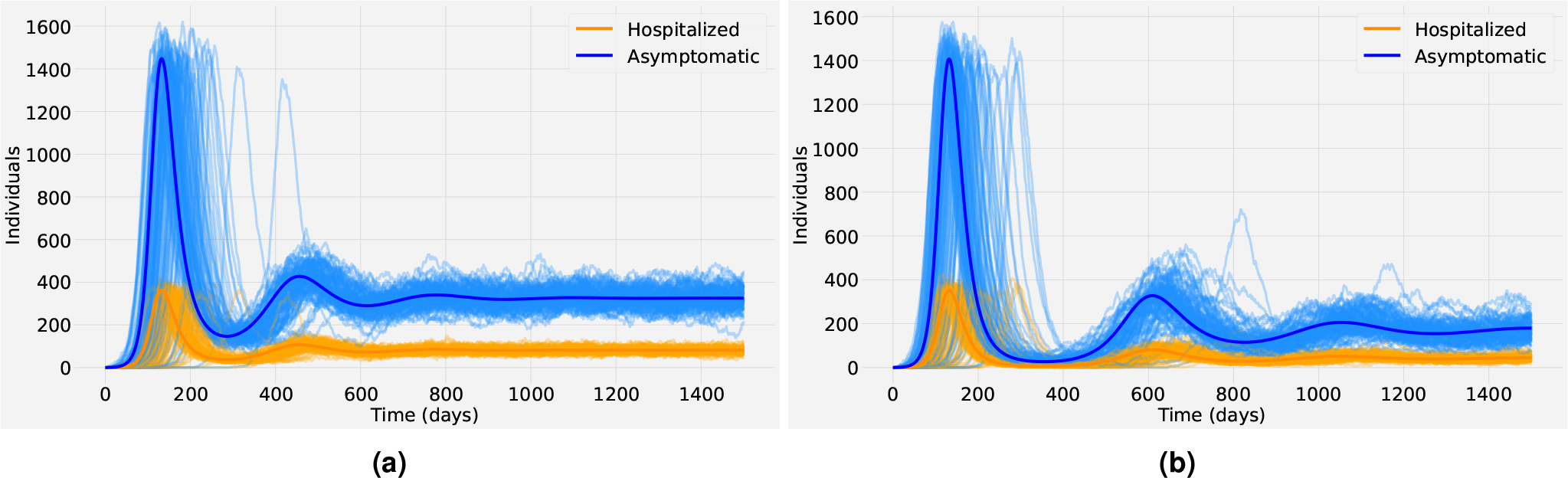
Stochastic realizations and the analytic mean-field solution for the asymptomatic class (blue) and the hospitalized class (orange). The basic reproduction number for the human population corresponds to a supercritical regime 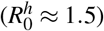 and the spillover rate is assumed to be *τ* = 10^*-*5^. In (a) the average duration of natural immunity is 180 days and in (b) is 360 days. Other parameters are as shown in Table 1 in the SM.

### 2.3 The 2022 monkeypox outbreak outside endemic regions: a case study with empirical data

While we argue that COVID-19 dynamics could reassemble the scenario discussed in Fig. 6b, with up to 60,000 estimated cases occurring in the Basque Country, Spain (2.2 *·* 10^6^ inhabitants) during the first few weeks of establishment, the available data is limited and already showing an exponential growth that was quickly interrupted by the extreme lockdown measures. Thus, it is not possible to investigate quantitatively well the initial phase of COVID-19. However, models can bridge the missing gaps in epidemiological dynamics, providing possible scenarios of disease spreading and quantifying the uncertainties resulting from the missing data. On the other hand, for the monkeypox outbreaks, no extreme human intervention took place, and hence the available data describe better an unperturbed epidemiological situation.

We present weekly confirmed cases from the start of the outbreak in the four most affected European countries (Spain, France, Germany, and the United Kingdom) until 23 September 2022 (see Figure 7). The incidence time series and confirmed cases for these and several other countries can be obtained from an open-access database presented in Kraemer et al.^40^. In the case of the European Region data is also available via the European Center for Disease Control (ECDC) and the WHO Regional Office for Europe through The European Surveillance System (TESSy)^41^. For each country, the data is represented as bar plots. We remark that given the unexpected outbreak and the large geographical spread of the disease, the actual number of cases is likely to be underestimated and the extent of asymptomatic infection is unknown. Hence, the stochastic SI-SHAR model is calibrated to the data for each country via the hospitalized class *H* used as a proxy for cases showing severe disease symptoms which are easily detected. The mean-field analytical solution of the SI-SHAR model together with an ensemble of stochastic realizations can successfully reproduce the epidemiological dynamics of these monkeypox outbreaks (see Figure 7). The data presented in Figure 7 suggest that the current outbreaks present stage III dynamics of pathogen emergence, and differently from COVID-19 (expected to present stage IV dynamics of pathogen emergence), the possibility of a huge pandemic is very low. Although confirmed monkeypox cases decreased significantly in the European region, further strategies toward elimination are essential to avoid subsequent evolution of the monkeypox virus that can result in higher transmission^42^. Note that while this decline might be attributed to public health interventions and behavioral changes in the population due to increased risk perception, no drastic intervention measures were needed to control the outbreaks, and that can be considered as an indicator of low pathogen virulence, i.e., with low human-to-human transmission rate (much lower than the SARS-CoV-2 viruses).

**Figure 7.**
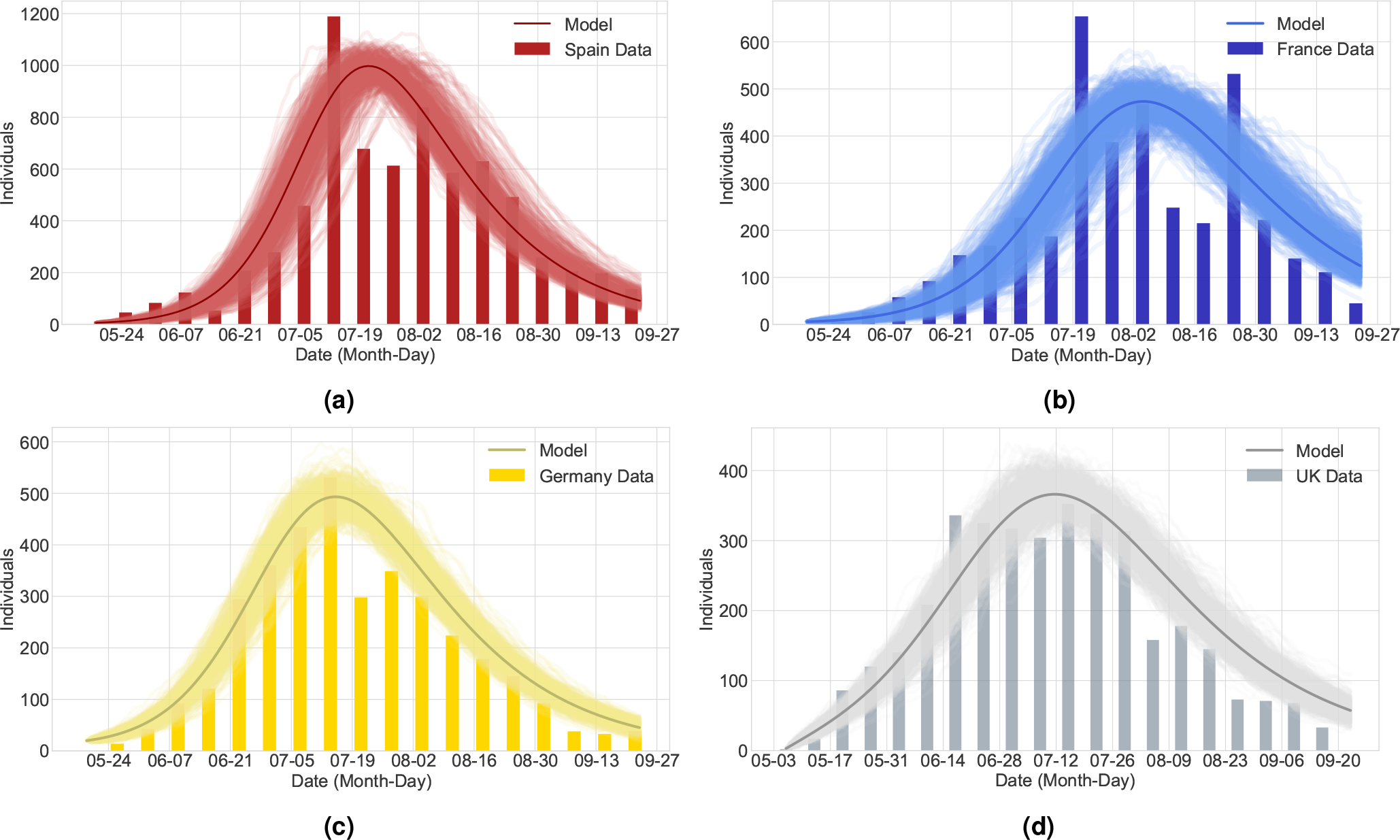
Weekly laboratory-confirmed monkeypox cases in (a) Spain, (b) France, (c) Germany, and (d) the United Kingdom, from May to September 2022. The data is represented by bar plots. The stochastic SI-SHAR model is calibrated to the data for each country via the hospitalized class *H* used as a proxy for cases showing severe disease symptoms. The mean-field solution (solid line) together with an ensemble of 500 stochastic realizations is shown for each country.

## 3 Discussion

The emergence of environmental and wildlife pathogens in human populations is a process that has occurred several times and is expected to keep happening in the near future. The most alarming part is that significant evidence indicates that zoonotic pandemics are becoming more frequent and rising in cost^6,9^. Current global disease control to address future pandemics focuses mainly on pandemic preparedness and response, such as vaccine and drug development, and healthcare system strengthening. Although these actions are crucial and have a useful effect, they overlook the critical need to prevent pandemics before they are established^6^. Since the natural reservoir is often unknown, control strategies directed at spillover prevention should focus on reducing activities that are expected to increase the overall risk of zoonotic disease outbreaks^7^. Multiple studies suggest that ecological disruption, including deforestation, forest degradation, habitat fragmentation, agricultural expansion, wildlife trading, and climate change, results in increasing levels of pathogen exposure and created conditions for successful spillover events^1–3^. Hence, we cannot expect a decrease in the risk of newly emerging diseases unless we address ecological degradation, population growth, and climate change.

In this work, we have proposed a minimalistic modeling framework to describe the spillover interface and explore conditions that lead to disease emergence in human populations. In contrast to current mathematical models of the zoonotic spillover interface, our model explicitly considers the dynamics in the non-human species and provides a stochastic treatment of spillover dynamics. Yet, the model is parsimonious and essentially mimics the stochastic dynamics of two SIR-type processes connected via spillover events. Through several numerical experiments, we illustrated the role of the reservoir dynamics, the host-specific reproduction numbers, and the spillover transmission rate on the global epidemiological dynamics of the system. The model shows that if the human epidemic is within a subcritical regime, spillover events can significantly affect prevalence levels in the human host. Particularly, a high spillover rate can cause isolated outbreaks, sometimes of significant sizes, which lead to a positive stationary number of new infections in the mean-field approximation, even when large stochastic fluctuations are detected. In other words, although most outbreaks are caused by a new pathogen with pandemic potential, in nature, most of these outbreaks usually die out due to the stochastic nature of the process. Nevertheless, the presence of spillover events coming from outside the human population can avoid long-lasting extinction and lead to new and frequently unexpected large outbreaks occurring at different points in time in different regions of the world, as was observed for COVID-19 epidemics. These fluctuations, rather than details of the infection process, dominate the dynamical behavior of the system once the host-to-host transmission of a new pathogen can be maintained. Thus, neglecting critical stochastic fluctuations coming from cross-species transmission could lead to inaccurate epidemiological dynamics and flawed control interventions in the human population.

While there is no good quality data collected during the initial phase of COVID-19 epidemics, we have used numerical simulations to illustrate the impact of spillover events on the emergence and spread of new infectious diseases. As a case study, in this paper, we evaluate the 2022 monkeypox outbreak outside endemic regions, particularly within the European region as an example of a zoonotic disease driven by both person-to-person and reservoir-to-human transmission. Our results suggest that, differently from the observed COVID-19 dynamics, the 2022 monkeypox outbreaks do not represent a high public health risk for a pandemic, presenting limited transmission chains of human-to-human secondary infection. As a final remark, our modeling framework is a versatile tool that not only can be used to describe the general characteristics of the spillover interface but also can be easily adapted to build more complex models suitable for investigating important aspects of the disease emergence process for specific zoonotic pathogens.

## 4 Methods

The results presented here are based on a two-host stochastic Kermack-McKendrick-type system that includes the natural reservoir *N*_*r*_ and the human population *N*. After spillover events, pathogens can lead to disease in novel non-natural hosts. Nevertheless, in most cases, these pathogens cause only mild or asymptomatic infections which persist for a prolonged time in their reservoir hosts^24^. Hence, we assume an *S*_*r*_*I*_*r*_ (Susceptible-Infected) structure for the transmission dynamics in the reservoir population coupled to a *SHAR* (Susceptible-Hospitalized-Asymptomatic-Recovered) structure for the dynamics in the human population.

### 4.1 The SI-SHAR model

The reservoir model dynamics are described by the following reaction scheme

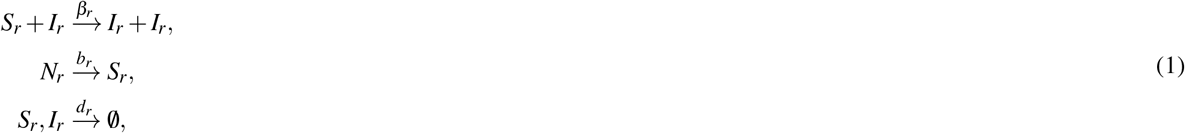

whereas the reaction scheme for human epidemiological dynamics is

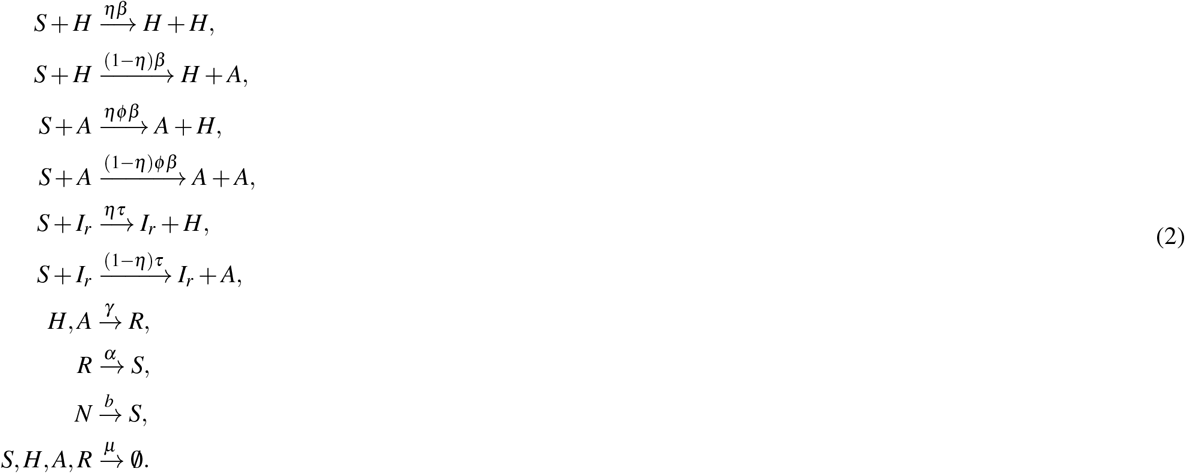

The SI-type model describes the dynamics of the reservoir population. We assume that natural animal host usually carries pathogens without presenting any clinical symptoms. While the pathogen transmission is well maintained in the population in stationary, the basic reproduction number in the reservoir set to in the supercritical regime, i.e. above one 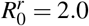 (with the average reservoir life expectancy assumed to be 1*/d*_*r*_ = 1 year, and the transmission rate *β*_*r*_ = 2*d*_*r*_). For simplicity, reservoirs can not recover from the infection, contributing to the force of infection throughout their life span.

The SHAR model is an extension of the well-known SIR framework in which the I class is divided into two groups labeled *H* and *A*, respectively: *H* stands for individuals developing a severe form of the disease and likely being hospitalized, while *A* refers to infected individuals who are asymptomatic or have a mild form of the disease. Besides the transmission rate in the human host *β*, the recovery rate *γ*, and the waning immunity rate *α* (already present in the general SIR formulation), the SHAR model includes two extra epidemiological parameters: the severity ratio *η*, that gives the fraction of infected individuals who develop severe symptoms, and the scaling factor *ϕ*, which differentiates the infectivity *ϕβ* of mild/asymptomatic infections with respect to the baseline infectivity *β* of severe/hospitalized cases^33^. The value of *ϕ* can be tuned to describe different situations: a value of *ϕ <* 1 indicates that severe cases have larger infectivity than mild cases (which in the case of infectious respiratory diseases, for example, could be linked to enhanced coughing and sneezing), while *ϕ >* 1 would be used to describe the scenario in which asymptomatic individuals and mild cases contribute more to the spread of the infection (e.g., due to their higher mobility and possibility of interaction) than severe cases (that are more likely to be detected and isolated). Here, the spillover factor *τ* refers to the possibility of susceptible individuals acquiring the disease via a spillover event, and couples the dynamics of the animal reservoir and the human hosts.

The spillover force of infection depends on the prevalence in the reservoir *I*_*r*_*/N*_*r*_ and the composite parameter *τ* = *cp*, which is the product of the reservoir-human contact rate *c* and the probability of infection given a contact *p*^10^. Infected human individuals in both the asymptomatic and hospitalized classes recover naturally at a rate *γ*, yet they can become susceptible again due to waning of immunity or even due to new virus variants at a rate *α*. Demographic dynamics are included in the reservoir via a birth rate *b*_*r*_ proportional to the reservoir total population *N*_*r*_ and the reservoir death rate *d*_*r*_. Likewise, humans are recruited into their susceptible class with a birth rate *b* proportional to the human population *N* and suffer from natural death at a rate *μ*.

Assuming that both the reservoir and the human population are in demographic equilibrium, the dynamics of the stochastic process are governed by the following master equation for the dynamics of the rate of change of states probabilities^33,34,43^

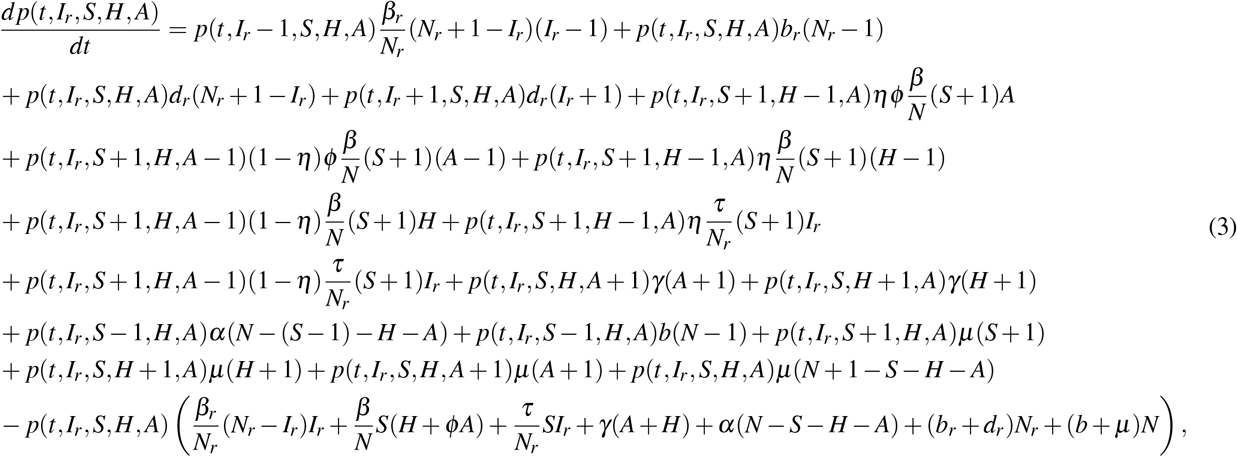

where *S*_*r*_ = *N*_*r*_ − *I*_*r*_ and *R* = *N* − (*S* + *H* + *A*). From the two-host Kermack-McKendrick-type model given as the master equation (3) we can obtain realizations of the stochastic process via the classical Doob-Gillespie direct method^44^ (an exact Monte Carlo method also known as the stochastic simulation algorithm (SSA)). The algorithm is implemented in StochSS^45^. Furthermore, defining the mean values and their derivatives

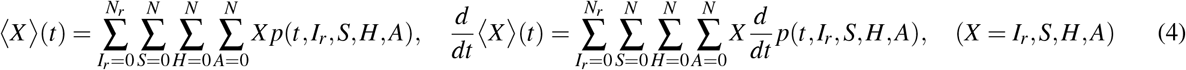

we obtain the mean-field approximation, i.e. neglecting variances and co-variance, the following dynamical systems for the mean values where we have omitted the ⟨*·*⟩ for notation simplification.

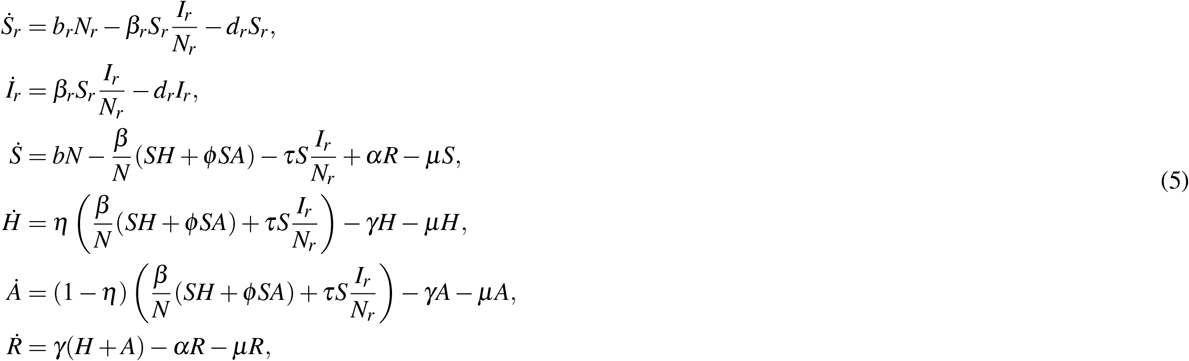

For a more detailed description of the mean-field approximation, please refer to^33,34^ and references therein. For simplicity, the equality of the birth and death rates is assumed for both species, respectively, so the system is at demographic equilibrium. For the mean-field model, the next-generation method^46^ can be used to obtain the following human basic reproduction number

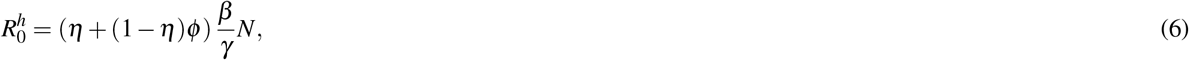

therefore for fixed *N, η, ϕ, γ*, the value of the human transmission rate *β* fully determines 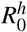 and vice-versa.

## Data availability

The datasets analyzed during the current study are available from the European Center for Disease Control (ECDC)^41^ and the open-access database presented in Kraemer et al.^40^

## Acknowledgements

This research is supported by the Basque Government through the “Mathematical Modeling Applied to Health” Project, BERC 2022-2025 program and by the Spanish Ministry of Sciences, Innovation and Universities: BCAM Severo Ochoa accreditation CEX2021-001142-S / MICIN / AEI / 10.13039/501100011033

## Author contributions statement

F.S, N.S., and M. A conceived the study. F.S wrote the original draft and performed the numerical simulations. N.S, J.B.V and M.A, developed the models and contributed to manuscript writing and critical discussions. M.A. supervised the project. All authors reviewed the manuscript.

## Additional information

The authors declare no competing interests. Correspondence and requests for materials should be addressed to M.A.

