## Supplementary material for "Modeling spillover dynamics: understanding emerging pathogens of public health concern": Suplementary Materials

October 2, 2023

#### S1 Mathematical analysis

Here, we present the stability analysis for the mean-field approximation of the two-host compartmental model given by the following system of nonlinear differential equations:

$$\begin{aligned}
\dot{S}_r &= b_r N_r - \beta_r S_r \frac{I_r}{N_r} - d_r S_r,\\
\dot{I}_r &= \beta_r S_r \frac{I_r}{N_r} - d_r I_r,\\
\dot{S} &= bN - \frac{\beta}{N}(SH + \phi SA) - \tau S \frac{I_r}{N_r} + \alpha R - \mu S,\\
\dot{H} &= \eta\left(\frac{\beta}{N}(SH + \phi SA) + \tau S \frac{I_r}{N_r}\right) - \gamma H - \mu H,\\
\dot{A} &= (1-\eta)\left(\frac{\beta}{N}(SH + \phi SA) + \tau S \frac{I_r}{N_r}\right) - \gamma A - \mu A,\\
\dot{R} &= \gamma(H + A) - \alpha R - \mu R.
\end{aligned} \tag{S1}$$

As described in the main text, we assume the equality of the birth and death rates, so both the reservoir and the human population are in demographic equilibrium. In other words, the total population is constant, hence, $N_r(t) = N_r^* > 0$ and $N(t) = N^* > 0$ for all $t > 0$.

The basic reproduction number for the reservoir population is given by $R_0^r = \beta_r/d_r$. The dynamics of the one-host deterministic SI reservoir model with standard incidence are well-understood (Saldaña and Velasco-Hernández, 2021; Vargas-De-León, 2011). The model has a disease-free equilibrium (DFE) and a unique endemic equilibrium. The basic reproduction number $R_0^r$ has the threshold property that determines the local (Van den Driessche and Watmough, 2002) and global (Vargas-De-León, 2011) stability of the equilibria. By definition, the reservoir population can maintain the infection at an endemic level (Alexander et al., 2018), therefore, we assume $R_0^r > 1$ so the endemic equilibrium (EE) for the reservoir system given by

$$I_r^* = (1 - 1/R_0^r)N_r^*, \quad S_r^* = N_r^* - I_r^*, \tag{S2}$$

and is asymptotically stable. The basic reproduction number for the human population in isolation (disregarding spillover infections) is given by the following expression

$$R_0^h = \frac{(\eta + (1-\eta)\phi)\beta}{(\gamma + \mu)}. \tag{S3}$$

The first term in (S3) measures the contribution of the hospitalized class $H$ to the force of infection, whereas the second term quantifies the contribution of the asymptomatic class $A$.

To obtain the global basic reproduction number, we use the next-generation operator (Diekmann et al., 1990) and the method of (Van den Driessche and Watmough, 2002) obtaining the following next-generation matrix

$$\mathbf{K} = \begin{bmatrix} R_0^r & 0 & 0 \\ c_1 & \dfrac{\eta\beta}{\gamma+\mu} & \dfrac{\eta\phi\beta}{\gamma+\mu} \\ c_2 & \dfrac{(1-\eta)\beta}{\gamma+\mu} & \dfrac{(1-\eta)\phi\beta}{\gamma+\mu} \end{bmatrix} \tag{S4}$$

where $c_1$, $c_2$ are positive constants. The global basic reproduction number $R_0$ of the two-host SI-SHARS process (S1) is defined as the spectral radius of $\mathbf{K}$ and is given by

$$R_0 = \max\{R_0^r, R_0^h\}. \tag{S5}$$

It follows that $R_0^r > 1$ implies $R_0 > 1$. Furthermore, in a deterministic setting, for the coupled SI-SHARS model (S1), if the reservoir is at the EE and the spillover rate satisfies $\tau > 0$, then no DFE exists for the human population. This result can be easily shown from the human equations for equilibria given by

$$0 = bN - \frac{\beta}{N}(SH + \phi SA) - \tau S\frac{I_r^*}{N_r^*} + \alpha R - \mu S, \tag{S6}$$

$$0 = \eta\left(\frac{\beta}{N}(SH + \phi SA) + \tau S\frac{I_r^*}{N_r^*}\right) - \gamma H - \mu H, \tag{S7}$$

$$0 = (1-\eta)\left(\frac{\beta}{N}(SH + \phi SA) + \tau S\frac{I_r^*}{N_r^*}\right) - \gamma A - \mu A, \tag{S8}$$

$$0 = \gamma(H + A) - \alpha R - \mu R, \tag{S9}$$

If the human population is at the DFE i.e $H^* = A^* = 0$, then (S9) implies $R^* = 0$, and (S7) implies $S^* = 0$, therefore $N = 0$ which is not biologically feasible. To find an analytical expression for the EE observe that from equations (S7)-(S8) we have

$$\frac{\beta S^*}{N}(H^* + \phi A^* + \rho) = \frac{\gamma+\mu}{\eta}H^* = \frac{\gamma+\mu}{1-\eta}A^* \tag{S10}$$

where $\rho$ is a constant that satisfies $\rho = \tau I_r^* N/\beta N_r^*$. Therefore, we can obtain the asymptomatic class in terms of the hospitalized individuals as

$$A^* = \frac{1-\eta}{\eta}H^*. \tag{S11}$$

Likewise, using equations (S9) and (S11), we can obtain the equilibrium for the recovered class as

$$R^* = \frac{\gamma H^*}{\eta(\alpha+\mu)}. \tag{S12}$$

Now observe that $S^* = N - H^* - A^* - R^*$, therefore using the above results we obtain

$$S^* = N^* - \frac{\alpha+\mu+\gamma}{\eta(\alpha+\mu)}H^*. \tag{S13}$$

Substituting $S^*$, $A^*$, and $R^*$ into equation (S6) we get

$$bN - \frac{\beta}{N}\left(N - \frac{\alpha+\mu+\gamma}{\eta(\alpha+\mu)}H^*\right)\left(H^* + \frac{\phi(1-\eta)}{\eta}H^* + \rho + \mu\frac{N}{\beta}\right) + \frac{\alpha\gamma}{\eta(\alpha+\mu)}H^* = 0. \tag{S14}$$

The hospitalized class at equilibrium is therefore given by the solution of the following quadratic equation

$$a(H^*)^2 + bH^* + c = 0 \tag{S15}$$

where

$$a = \frac{\beta(\alpha+\mu+\gamma)}{N\eta(\alpha+\mu)}\left(1+\frac{\phi(1-\eta)}{\eta}\right), \tag{S16}$$

$$b = -\beta\left(1+\frac{\phi(1-\eta)}{\eta}\right)+\frac{\beta(\alpha+\mu+\gamma)}{N\eta(\alpha+\mu)}\left(\rho+\mu\frac{N}{\beta}\right)+\frac{\alpha\gamma}{\eta(\alpha+\mu)}, \tag{S17}$$

$$c = -\beta\rho. \tag{S18}$$

Observe that $a > 0$ and $c < 0$, hence $-4ac > 0$ and $\sqrt{b^2-4ac} > |b|$, therefore the only biologically feasible equilibrium is

$$H^* = \frac{-b+\sqrt{b^2-4ac}}{2a}. \tag{S19}$$

Note that the unique EE for the human SHAR model $(S^*, H^*, A^*, R^*)$ always exists if $R_0^r > 1$. This result is valid even if the basic reproduction number for the human population $R_0^h$ is less than one. From the analysis in this section and Theorem 2 in (Van den Driessche and Watmough, 2002), we obtain the following result.

**Theorem 1.** *If the basic reproduction number for the reservoir population satisfies $R_0^r > 1$, then the SI-SHAR model has a unique endemic equilibrium given by $(S_r^*, I_r^*, S^*, H^*, A^*, R^*)$ which is locally asymptotically stable.*

#### S2 Effect of the reservoir prevalence on the human outbreaks

A reservoir can be simply defined as an animal population that can maintain a virus in circulation (Wasik et al., 2019). A broader definition for reservoir can also be considered as one or more epidemiologically linked host species or environments in which a pathogen is self-sustained (Alexander et al., 2018). From these definitions is clear that at some point the natural reservoir will reach an endemic equilibrium. Nevertheless, one might argue that spillover events might occur, although with a lower probability, even if the reservoir is still far from endemic levels. Here, we consider such a scenario. Figure S1 corresponds to stage II of pathogen emergence, hence, the outbreak in the human population is fully driven by spillover events and no further human-to-human secondary transmission is possible. Figure S2 corresponds to stage III of pathogen emergence, hence, human-to-human secondary infections are possible but they cannot lead by themselves (without further spillover events) to a large outbreak in the human population. Stage IV is represented in Figure S3. Unlike Stages II-III which correspond to subcritical threshold regimes, Stage IV corresponds to a supercritical regime that leads to an exponential growth of infection (see Figure S3).

Comparing the epidemiological dynamics observed in Figures S1-S3 with their counterpart presented in the main text, it is evident that no significant qualitative changes are presented. However, some quantitative differences appear. For example, on average, the stochastic realizations presented here present greater variance than the ones in the main text. In other words, considering the early outbreak dynamics in the animal reservoir might lead to bigger stochastic fluctuations in the human host. Furthermore, for Stages II and III (see Figures S1-S2) the epidemic curve follows a more symmetric pattern in comparison with its counterpart in the main text. This behavior is a consequence of a lower spillover force of infection that depends on the prevalence in the reservoir $I_r/N_r$ and the composite parameter $\tau = cp$, defined as the product of the reservoir-human contact rate $c$ and the probability of infection given a contact $p$. In the case explored in the main text, the animal reservoir is close to endemic levels $I_r \approx I_r^*$, where $I_r^*$ is the EE. Here, we are considering $I_r \ll I_r^*$ for early times but $I_r$ converges asymptotically to $I_r^*$. Hence, before the animal reservoir reaches prevalence levels close to the EE, the spillover force of infection is low and spillover events are scarce. If human epidemiological dynamics are within a subcritical regime and the spillover force of infection is low, then human incidence grows slowly and the epidemic curve takes its classical symmetric form (e.g. Figures S2). For a supercritical regime, that is, when the human basic reproduction number is above 1, the spillover force of infection no longer plays a vital role in human outbreaks. In this case, even with a few spillover events, human-to-human secondary infections lead to exponential epidemic growth

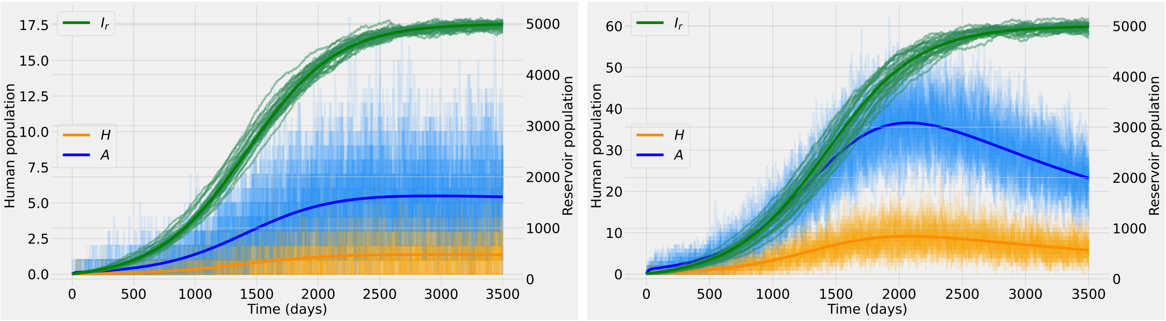


(a) (b)

Figure S1: Stochastic realizations and the analytic mean-field solution for the asymptomatic class (blue), the hospitalized class (orange), and the infected class for the animal reservoir (green). The dynamics correspond to stage II of pathogen emergence, so no further human-to-human secondary transmission is possible, $\beta = 0$ and therefore $R_0^h = 0$. In (a) the spillover rate is $\tau = 10^{-4}$ and in (b) is $\tau = 10^{-3}$.

(see Figure S3). Furthermore, under these conditions, the epidemic outbreak in the human population arises long before the reservoir prevalence reaches endemic levels (see Figure S3).

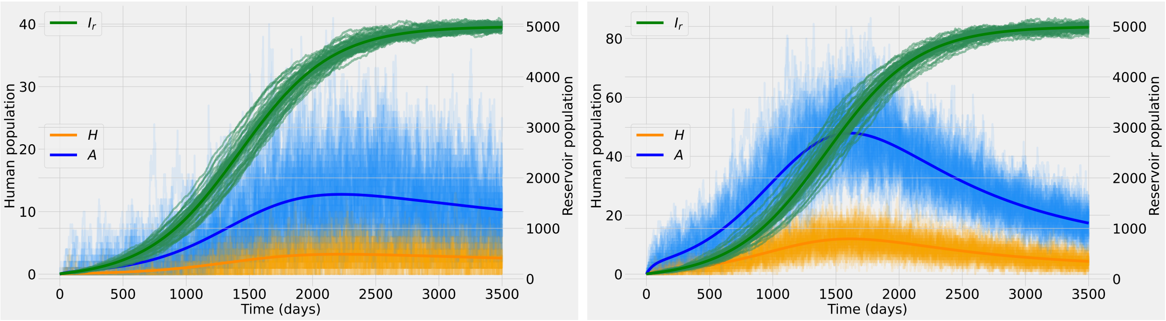


(a) (b)

Figure S2: Stochastic realizations and the analytic mean-field solution for the asymptomatic class (blue), the hospitalized class (orange), and the infected class for the animal reservoir (green). The dynamics correspond to stage III of pathogen emergence, in particular, $0 < R_0^h \approx 0.9 < 1$. In (a) the spillover rate is $\tau = 10^{-4}$ and in (b) is $\tau = 10^{-3}$.

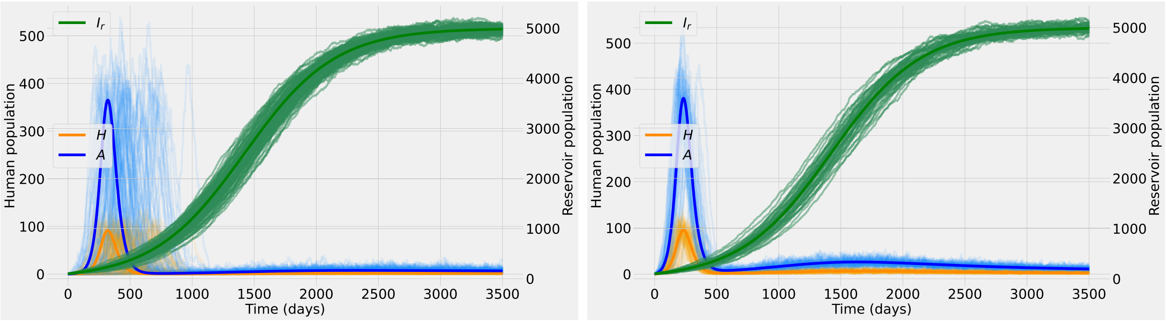


(a) (b)

Figure S3: Stochastic realizations and the analytic mean-field solution for the asymptomatic class (blue), the hospitalized class (orange), and the infected class for the animal reservoir (green). The dynamics correspond to stage IV of pathogen emergence with $1 < R_0^h \approx 1.4$. In (a) the spillover rate is $\tau = 10^{-4}$ and in (b) is $\tau = 10^{-3}$.

### S3 Model parameters

The parameters used in the numerical experiments are summarized below.

Table S1: Baseline model parameters. Other parameter values eventually used for numerical simulations will be explicitly stated in figure captions. The estimations are taken from previous studies on COVID-19 Aguiar et al. (2021, 2020a); Srivasrav et al. (2022); Aguiar et al. (2022, 2020b); Aguiar and Stollenwerk (2020); Chen et al. (2020) except $\tau$ for which there is no data, thus we assume arbitrary values for exploratory investigation. The total population for both reservoir and human population is assumed to be $10,000$.

| Parameter | Range | Mean |
|---|---|---|
| $1/d_r$ (reservoir life expectancy) | $[0, 10]$ years | 1 years |
| $\beta_r$ (reservoir transmission rate) | $[0, 20]$ years$^{-1}$ | 2 years$^{-1}$ |
| $1/\mu$ (human life expectancy) | $[0, 90]$ years | 60 years |
| $1/\gamma$ (human mean infectious period) | $[0, 30]$ days | 15 days |
| $\beta$ (human transmission rate) | $[0, 4/15]$ days$^{-1}$ | 1/15 days$^{-1}$ |
| $\eta$ (fraction that develop severe disease) | $[0, 1]$ | 0.2 |
| $\phi$ (change of infectivity of $A$ versus the infectivity $H$) | $[0, 5]$ | 1.5 |
| $1/\alpha$ (human mean duration of natural immunity) | $[0, 360]$ days | 180 days |
| $\tau$ (spillover transmission rate) | $[1e^{-6}, 1e^{-3}]$ days$^{-1}$ | $1e^{-5}$ days$^{-1}$ |
